# The Economic Burden of Dengue: A Systematic Literature Review of Cost-of-Illness Studies

**DOI:** 10.1101/2025.08.21.25334162

**Authors:** Doungporn Leelavanich, Ilaria Dorigatti, Hugo C. Turner

## Abstract

**Background:** Dengue, a vector-borne disease caused by the dengue virus, has emerged as a global public health concern, given the tenfold rise in reported cases over the last two decades. In light of the upcoming dengue interventions, country-specific cost-of-illness estimates are required to evaluate the cost-effectiveness of new interventions against dengue. This study aims to conduct an updated systematic review of dengue cost-of-illness studies, extracting the relevant data, and conducting regression analysis to explore potential factors contributing to the cost variations among countries. We used the MEDLINE, EMBASE, PubMed, and Web of Science databases to systematically search for published dengue cost-of-illness studies reporting primary costs per dengue case. A descriptive analysis was conducted across all extracted studies. Linear regression analysis was performed to investigate the association between the GDP per capita and cost per case. The quality of the included studies was also assessed. Fifty-six studies were included, of which 22 used the societal perspective. The reported total cost per case ranged from $15.0 for outpatients in Burkina Faso to $9,386.1 for intensive care unit patients in Mexico. Linear regression analysis revealed that the cost of dengue illness varies significantly across countries and regions, and was positively related to the setting’s GDP per capita. The quality assessment demonstrated that improvements are needed in future studies, particularly in the reporting of the methodology. Future research should focus on understanding other drivers of cost variations beyond GDP per capita to improve the cost estimates for economic evaluation studies. The results presented in this study can serve as crucial input parameters for future economic evaluations, supporting decision makers in allocating resources for dengue intervention programs.

## Introduction

Dengue is a growing global public health concern, particularly affecting populations in tropical and subtropical regions (1). It is a vector-borne disease caused by the dengue virus (DENV), which has four serotypes: DENV-1, DENV-2, DENV-3, and DENV-4 (1). The virus is transmitted to humans by mosquitoes, mainly *Aedes aegypti*, and to a lesser extent, *Aedes albopictus* (1). The majority of people infected with dengue are usually asymptomatic or have a self-limiting febrile illness (Dengue Fever (DF)) (1, 2). However, the disease can progress to more severe forms, including Dengue Haemorrhagic Fever (DHF) and Dengue Shock Syndrome (DSS), which can be life-threatening (1, 2).

According to the World Health Organization (WHO), dengue is a priority high-risk disease (3), and the number of reported cases increased tenfold over the last two decades, from 0.5 million in 2000 to 6.5 million in 2019 (1) The global economic burden of dengue was estimated to be $8.9 billion in 2013, with 42% resulting from productivity losses (4).

To date, there is no specific therapeutic treatment for dengue, making preventive interventions vital (1, 2). Standard vector control tools have been unable to sustainably control dengue, and a range of novel interventions are under development and becoming available (including vaccines and biocontrol strategies) (5). Economic evaluation studies offer a valuable tool to support decision-makers in determining which interventions could be cost-effective to implement, informing optimum resource allocation (6). Cost-of-illness studies (which estimate the economic burden of a specific disease or health condition on society) provide an essential input for such studies (6, 7). Numerous dengue cost-of-illness studies have been published to date, revealing significant variations in the costs of dengue across countries (7–9). This variation indicates that the economic burden of dengue is unique to each country, making it difficult to generalize the costs from one country to another. In 2013, Shepard *et al.* (10) found an association between the Gross Domestic Product (GDP) per capita and dengue related direct and productivity costs. However, these regression models only included data from 15 studies that were published before 2013. In addition, the direct costs considered were not stratified into direct medical and direct non-medical costs.

This study aims to conduct an updated systematic review of dengue cost-of-illness studies, extracting the relevant data, and conducting regression analysis to explore potential factors contributing to the cost variations among countries.

## Methods

### Screening and search strategy

A systematic literature review was conducted to gather dengue cost-of-illness data following the PRISMA 2020 guidelines (Appendix 1 in the supplementary file) (11). We searched for relevant articles from all country settings without time or language restrictions on May 22, 2024, across four databases: MEDLINE, EMBASE, Web of Science, and PubMed. Variants of the following search terms were used*: ‘(dengue OR DENV OR DENV-1 OR DENV-2 OR DENV-3 OR DENV-4) AND (cost of illness OR economic burden)’*. Details of the full search terms used for each database are provided in Appendix 2 in the supplementary file. The screening was performed by a single reviewer using the Covidence software. Titles and abstracts were screened for relevance to dengue cost-of-illness. Relevant articles were then retrieved for a full-text review. Articles were included if they met all of the following criteria: (i) the cost data were based on primary data collection and (ii) the cost per episode or cost per case was reported. Articles were excluded if they were conference abstracts, the estimates were not based on primary data, they did not report costs per case (i.e. reported total overall costs), the full texts were not available, or they were not written in English. Articles that focused on international travellers were also excluded, as these could skew the cost-of-illness data for the local population, which was the main focus of this study. Uncertainties regarding the inclusion or exclusion of certain studies were resolved through consultation with the co-authors.

### Data extraction and adjustment

The data extracted from each study included the publication year, country, costing approach, data collection approach, costing perspective, type of healthcare provider (public or private), treatment setting (outpatient, inpatient, and intensive care unit), and sample characteristics, such as case severity (DF or DHF/DSS) and age (child or adult).

In addition, we extracted the average cost per case data stratified by cost type (total costs, direct medical costs, direct non-medical costs, and productivity costs), either in the local currency or United States dollar (USD) if the local currency was not reported. To maximise the inclusion of extracted data in the analysis, several assumptions were made to maximise the inclusion of extracted data in the analysis, as outlined in Appendix 3 in the supplementary file.

As the cost data in different studies were collected in different years, it was necessary to adjust for inflation (12). All costs were converted and adjusted to 2023 USD values using the country’s GDP implicit price deflator, as recommended by Turner et al. (12). In brief, the cost data reported in USD were first converted to local currency using the exchange rate of the year reported in the articles. Then, local costs were adjusted for inflation using local inflation rates before being converted to 2023 USD values (12). All exchange rates and GDP deflator data were retrieved from the World Bank (13, 14). All data were recorded and adjusted using Microsoft® Excel.

### Data analysis

We first conducted a descriptive analysis of all the extracted studies. A quantitative analysis was then performed by calculating the average value of the extracted total cost per case, direct medical cost per case, direct non-medical cost per case, and productivity cost per case for each region and country. The WHO regional country groupings were used to classify countries, allowing comparison of average reported costs across regions (15). We included only countries with data on all cost subtypes for regional-level comparisons to ensure fair comparisons among regions (15). We also reported and compared the cost per case between public vs private settings; as well as costs for outpatients, inpatients, and critically ill patients treated in intensive care units (ICU patients) sub-groups, across countries. Cost data specifically related to critically ill ICU patients were considered separately from the inpatient sub-group.

In addition, we used linear regression to analyse the trends of the total cost per case, direct medical costs per case, direct non-medical costs per case, and productivity costs per case of outpatients and inpatients treated in public settings to understand the factors driving the variation in each of the cost types. The total costs were defined as the sum of the total direct medical costs (the sum of direct medical costs borne by both the government and patients), the total direct non-medical costs (the sum of direct non-medical costs borne by both the government and patients), and productivity costs. Studies that reported costs borne solely by patients or solely by the government were excluded from the regression analysis for the direct medical costs.

A regression analysis for the costs related to treatment in private facilities and ICU patients was not conducted owing to the limited available data. In addition, we used the World Bank’s latest GDP per capita as a proxy to represent the economic status of each country. We used a log-log transformation in the linear regression model, i.e. used the logarithm of cost as the dependent variable and the logarithm of GDP per capita as the independent variable as outlined below:

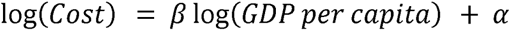

Where:

– *log(Cost)* is the logarithm of cost per case
– *β* is the coefficient
– *log(GDP per capita)* is the logarithm of GDP per capita
– *α* is the intercept

Appendix 4 in the supplementary file presents additional assumptions pertinent to the data analysis. In this study, linear regression was conducted in RStudio version 2023.09.1+494.

### Quality assessment

The quality of the included studies was assessed using a recently published cost-of-illness (COI) consensus-based checklist by Schnitzler et al. (16). This checklist includes 17 criteria, each graded on four levels: yes, partial, no, unclear, and not applicable. We assigned “yes” to studies that clearly described the criteria points, “no” to those that did not describe them at all and where it could not be inferred from the text, “unclear” if the points were implied but not explicitly described, and “partial” if the studies met some but not all parts of the criteria. “Not applicable” was used if the criterion did not apply to the study’s context.

## Results

A total of 1,986 studies were identified from the databases. After removing duplicates and screening titles and abstracts, 299 studies remained. Of these, 243 studies were excluded because they did not meet the eligibility criteria. Therefore, 56 studies were included in the final analysis. The detailed study selection process is shown in Fig. 1.

**Fig. 1.**
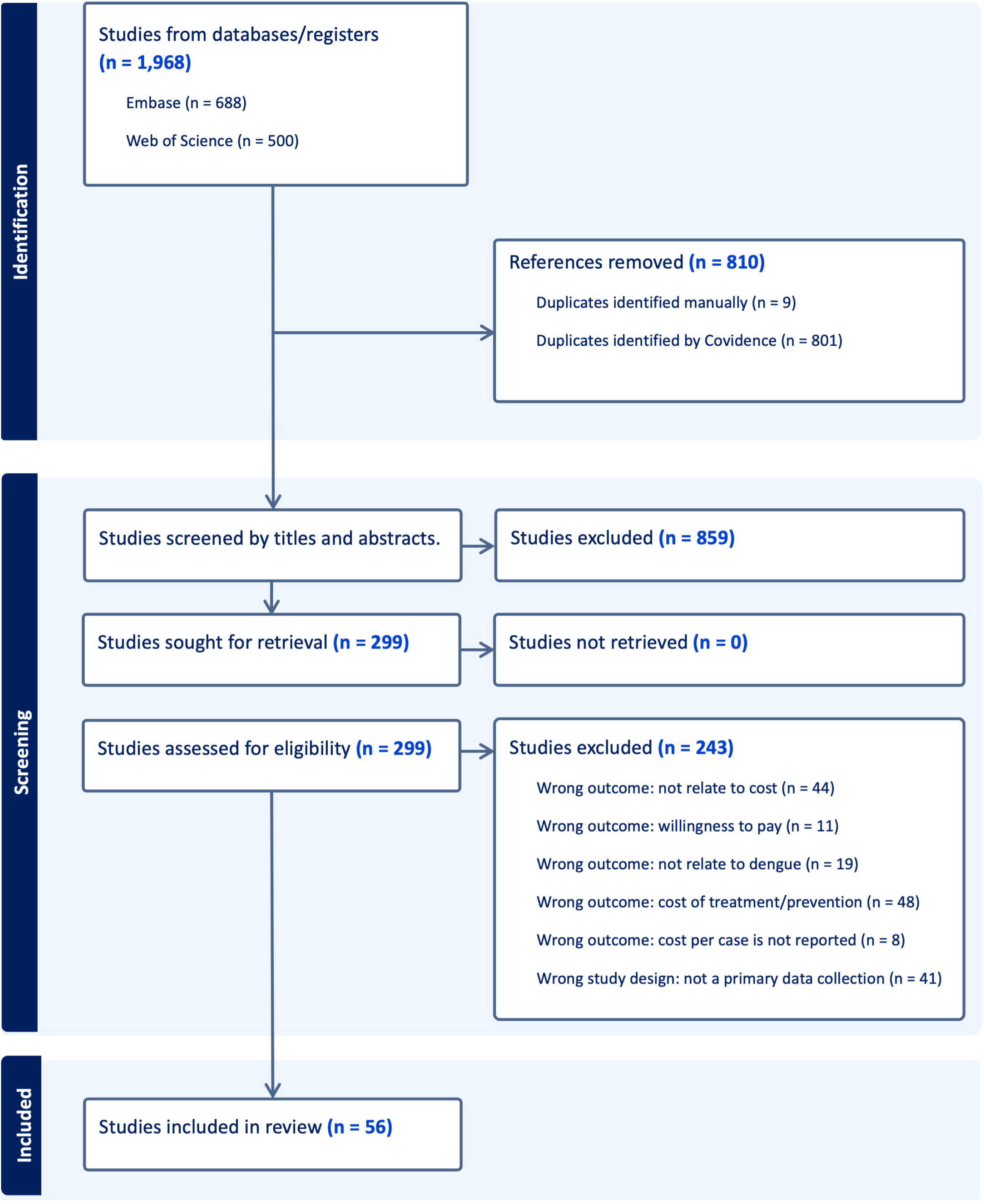
Prisma flow of the study selection

### Descriptive analysis of included studies

The descriptive analysis of the identified studies is summarised in Table 1. Cost-of-illness studies from 26 countries were captured, with several studies covering more than one country. The majority of the studies captured countries in Southeast Asia (24 studies) (9, 17–39). This was followed by the Western Pacific region (20 studies) (8, 29, 33, 36, 40–55). Only one study has focused on dengue in Africa (9). Viet Nam (8, 29, 33, 43, 45, 47, 49, 51, 54) and India (19, 20, 23, 25, 27, 31, 35, 37, 56) had the highest number of studies, with nine studies each. This was followed by Thailand (32, 33, 36, 38, 39), Cambodia (9, 36, 52, 53, 55), and Brazil (36, 57–60), each with five studies, while China (40–42, 46) and Sri Lanka (18, 21, 22, 61) had four studies each.

**Table 1.** Summary of the descriptive analysis.

| Categories | Number of studies | References |
| --- | --- | --- |
| **WHO regions** |  |  |
| South-east Asia | 24 | (9, 17-39) |
| Western Pacific | 20 | (8, 29, 33, 36, 40-55) |
| Americas | 12 | (33, 36, 57-60, 63-66, 68, 69) |
| Eastern Mediterranean | 4 | (30, 62, 67, 70) |
| African | 1 | (9) |
| **Perspective** |  |  |
| Societal | 22 | (9, 17, 18, 21, 24, 28, 31, 33, 36, 39, 41, 43, 44, 47, 48, 50, 56, 57, 62-65) |
| Household | 21 | (19, 23, 25, 26, 29, 30, 32, 34, 35, 38, 40, 45, 49, 51-55, 67-69) |
| Government | 5 | (22, 46, 59-61, 66) |
| **Private health insurance program** | 2 | (58, 59) |
| **Unclear** | 6 | (8, 20, 27, 37, 42, 70) |
| **Provider setting** |  |  |
| **Public** | 44 | (8, 9, 17-70) |
| **Private** | 17 | (17, 19, 26-28, 31, 35, 37, 50, 53, 55-59, 69, 70) |
| **Both public and private (overall)** | 7 | (20, 36, 48, 52, 55, 62, 64) |
| **Treatment setting** |  |  |
| **Outpatients** | 16 | (9, 24, 25, 28, 33, 36, 40, 44, 45, 50, 56, 57, 63, 65, 68, 69) |
| **Inpatients** | 44 | (8, 9, 17, 20-28, 30-33, 35-40, 42, 44-51, 53, 54, 56-58, 60, 61, 63, 65, 67-70) |
| **Intensive care unit** | 4 | (37, 43, 61, 65) |
| **Other** | 13 | (18, 19, 24, 29, 34, 41, 44, 52, 59, 62, 64, 66, 69) |
| **Unclear** | 1 | (55) |
| **Data collection approach** |  |  |
| **Retrospective** | 31 | (8, 23, 25-27, 31, 34, 38, 40-48, 50, 51, 55, 58-63, 65-69) |
| **Prospective** | 20 | (9, 18-22, 24, 29, 30, 32, 33, 35, 36, 39, 49, 52, 53, 57, 64, 70) |
| **Both Retrospective and prospective** | 3 | (17, 28, 56) |
| **Unclear** | 2 | (37, 54) |
| **Costing approach** |  |  |
| **Micro-costing** | 46 | (8, 9, 17, 19, 21-23, 25-35, 37-45, 47-49, 51-55, 57-59, 62, 63, 65-70) |
| **Macro-costing** | 7 | (18, 24, 36, 46, 50, 56, 61) |
| **Both micro-costing and macro-costing** | 1 | (64) |
| **Unclear** | 2 | (20, 60) |

Societal perspective was the most commonly used (22 studies) (9, 17, 18, 21, 24, 28, 31, 33, 36, 39, 41, 43, 44, 47, 48, 50, 56, 57, 62–65). Notably, among these 22 studies, only 14 reported all the types of costs. Two studies described their approach as a quasi-or restricted-societal perspective, which includes direct medical costs and productivity costs but excludes direct non-medical costs (19, 44).

The majority of the studies were conducted in a public healthcare provider setting (44 out of 56) (8, 9, 17–70), with 17 out of 56 studies capturing private hospital settings (17, 19, 26–28, 31, 35, 37, 50, 53, 55–59, 69, 70). In addition, it was found that 44 studies focused on inpatient groups, while 16 studies focused on outpatients. Only four studies specifically targeted ICU patients (37, 43, 61, 65). Interestingly, two studies examined costs in informal medical settings, such as self-medication at home or obtaining medication from a pharmacy (24, 69).

Most studies collected data retrospectively (31 studies), whereas 20 studies collected data prospectively. Three studies collected direct medical costs retrospectively, while prospective methods were used to collect direct non-medical costs and productivity costs (17, 28, 56). Regarding the costing approach, 46 studies used a micro-costing approach (8, 9, 17, 19, 21–23, 25–35, 37–45, 47–49, 51–55, 57–59, 62, 63, 65–70). Seven studies used a macro-costing approach to collect cost-of-illness data (18, 24, 36, 46, 50, 56, 61). One study used both micro-costing and macro-costing approaches to collect costs borne to households and administrative costs, respectively (64). Further details of the descriptive analysis of all included studies are reported in Appendix 5 in the supplementary file.

### Cost per case and cost drivers

#### Regional overview

The average reported cost per case of dengue varied significantly across regions (Fig. 2). It was observed that not only the total cost but also the sub-types of costs were lowest in the African region. Direct medical costs were found to be the main driver of the total cost in Africa, Southeast Asia, and the Western Pacific. On the other hand, the total costs in the Americas were primarily driven by productivity costs. It should be noted that no studies conducted in the Eastern Mediterranean region collected data on all cost types, resulting in no average cost per case for this region.

**Fig. 2.**
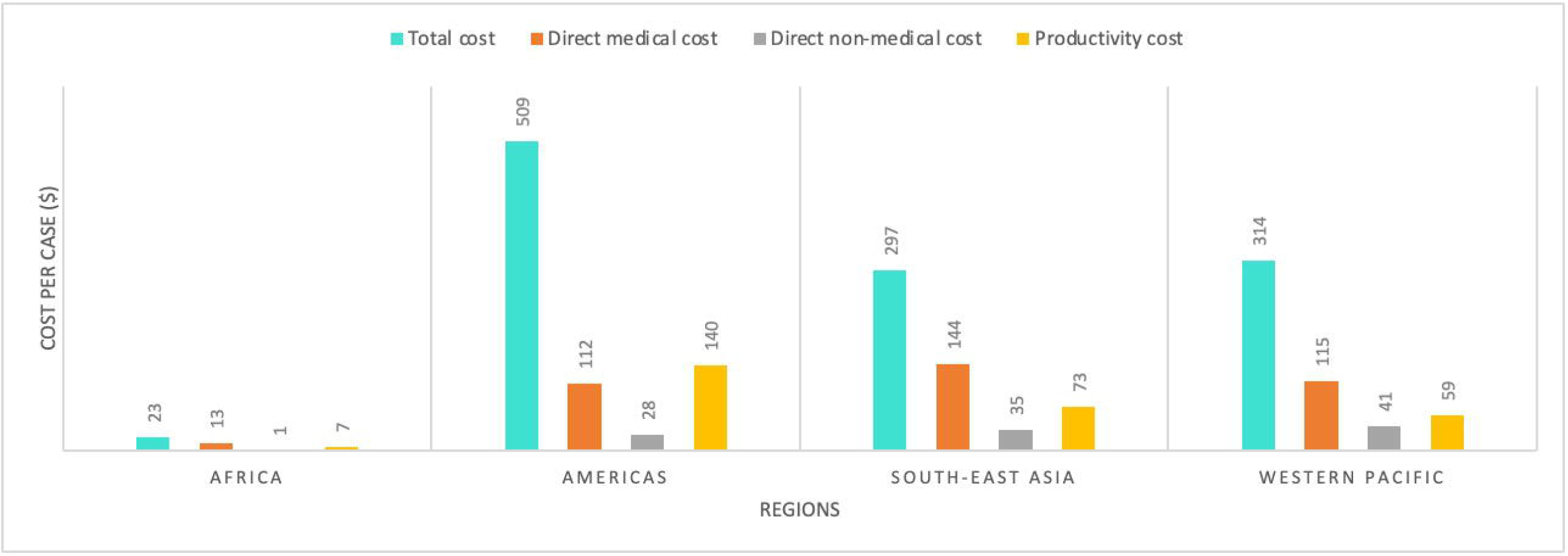
Average reported cost per case across regions

#### Cost per case in public settings

Across countries, the average total cost of an outpatient treated in public settings ranged from $15.0 in Burkina Faso to $128.0 in Colombia (Table 2). When considering only countries that reported all cost sub-types, the total costs were generally driven by productivity costs in most countries (Colombia, Thailand, Viet Nam). While direct medical costs had a greater influence on the total cost in Burkina Faso and Kenya.

**Table 2.**
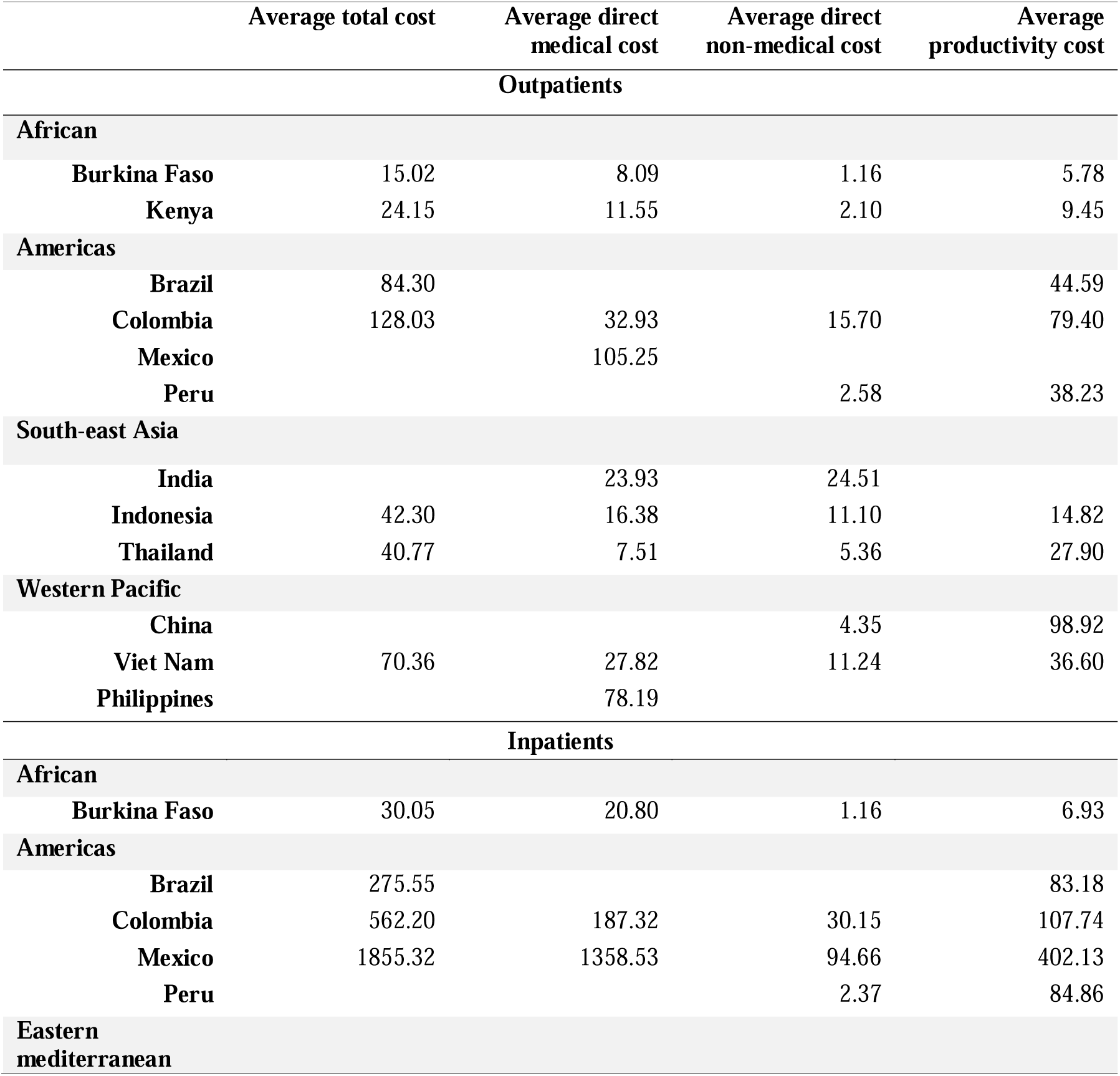
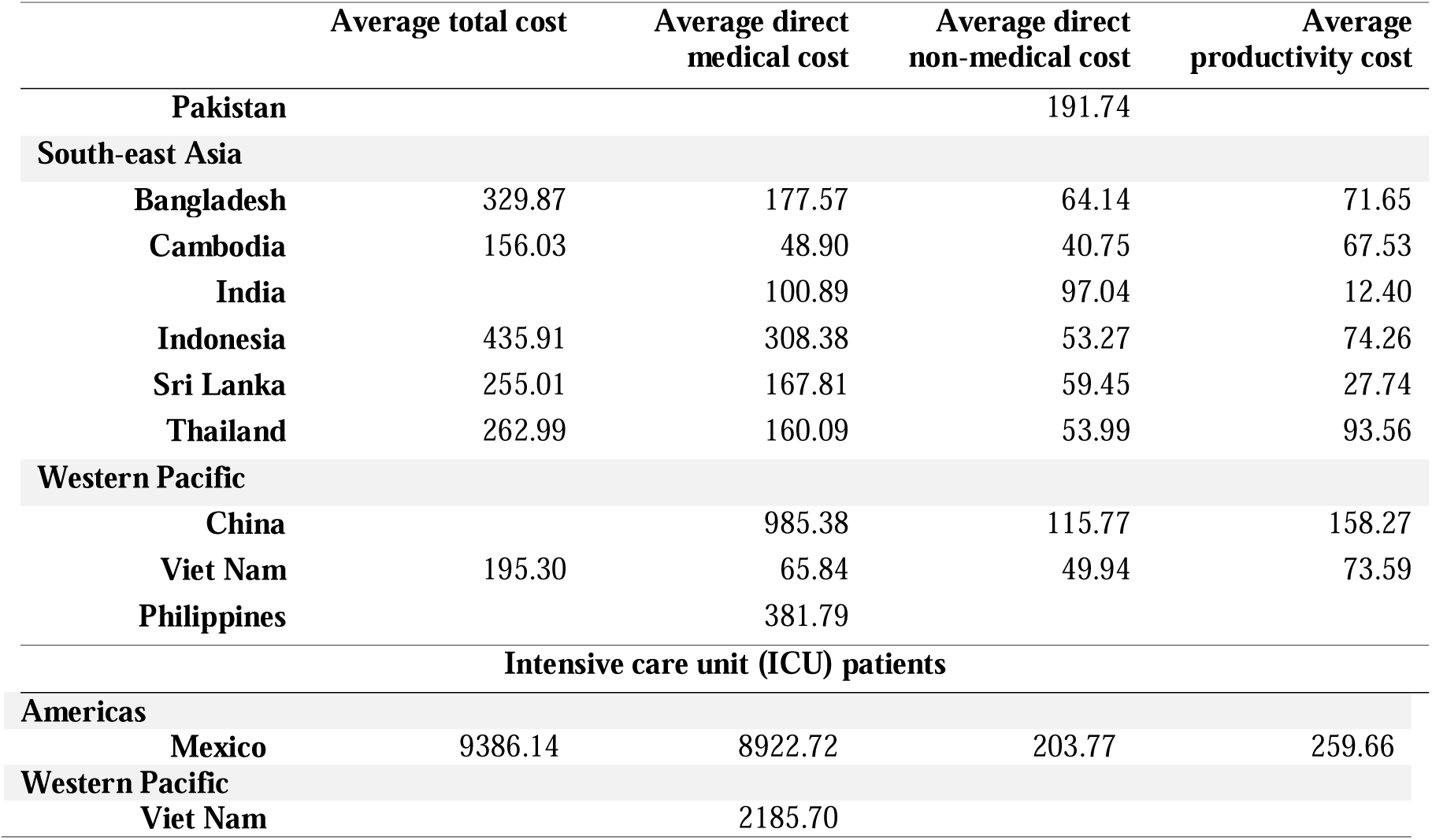
Average reported cost per case in public settings across countries (2023 USD)

|  | Average total cost | Average direct medical cost | Average direct non-medical cost | Average productivity cost |
| --- | --- | --- | --- | --- |
| **Outpatients** |  |  |  |  |
| **African** |  |  |  |  |
| Burkina Faso | 15.02 | 8.09 | 1.16 | 5.78 |
| Kenya | 24.15 | 11.55 | 2.10 | 9.45 |
| **Americas** |  |  |  |  |
| Brazil | 84.30 |  |  | 44.59 |
| Colombia | 128.03 | 32.93 | 15.70 | 79.40 |
| Mexico |  | 105.25 |  |  |
| Peru |  |  | 2.58 | 38.23 |
| **South-east Asia** |  |  |  |  |
| India |  | 23.93 | 24.51 |  |
| Indonesia | 42.30 | 16.38 | 11.10 | 14.82 |
| Thailand | 40.77 | 7.51 | 5.36 | 27.90 |
| **Western Pacific** |  |  |  |  |
| China |  |  | 4.35 | 98.92 |
| Viet Nam | 70.36 | 27.82 | 11.24 | 36.60 |
| Philippines |  | 78.19 |  |  |
| **Inpatients** |  |  |  |  |
| **African** |  |  |  |  |
| Burkina Faso | 30.05 | 20.80 | 1.16 | 6.93 |
| **Americas** |  |  |  |  |
| Brazil | 275.55 |  |  | 83.18 |
| Colombia | 562.20 | 187.32 | 30.15 | 107.74 |
| Mexico | 1855.32 | 1358.53 | 94.66 | 402.13 |
| Peru |  |  | 2.37 | 84.86 |
| **Eastern mediterranean** |  |  |  |  |
| **Pakistan** |  |  | 191.74 |  |
| **South-east Asia** |  |  |  |  |
| **Bangladesh** | 329.87 | 177.57 | 64.14 | 71.65 |
| **Cambodia** | 156.03 | 48.90 | 40.75 | 67.53 |
| **India** |  | 100.89 | 97.04 | 12.40 |
| **Indonesia** | 435.91 | 308.38 | 53.27 | 74.26 |
| **Sri Lanka** | 255.01 | 167.81 | 59.45 | 27.74 |
| **Thailand** | 262.99 | 160.09 | 53.99 | 93.56 |
| **Western Pacific** |  |  |  |  |
| **China** |  | 985.38 | 115.77 | 158.27 |
| **Viet Nam** | 195.30 | 65.84 | 49.94 | 73.59 |
| **Philippines** |  | 381.79 |  |  |
| **Intensive care unit (ICU) patients** |  |  |  |  |
| **Americas** |  |  |  |  |
| **Mexico** | 9386.14 | 8922.72 | 203.77 | 259.66 |
| **Western Pacific** |  |  |  |  |
| **Viet Nam** |  | 2185.70 |  |  |

The average total cost for inpatients treated in public settings ranged from $30.1 in Burkina Faso to $1,855.3 in Mexico (Table 2). Among countries that reported all cost sub-types, the costs were primarily driven by direct medical costs in most countries (Burkina Faso, Colombia, Mexico, Bangladesh, Indonesia, Sri Lanka, and Thailand). In contrast, productivity costs were the main driver only in Cambodia and Viet Nam.

Only Mexico and Viet Nam reported costs per case data for critically ill patients treated in ICUs within a public hospital (Table 2). The average direct medical costs for these cases were substantially higher than cases treated in outpatient and non-ICU inpatient settings ($8,922.7 for Mexico and $2,185.7 for Viet Nam). The direct medical costs were 85 times and 79 times higher than outpatient costs in Mexico and Viet Nam, respectively; and 7 times and 33 times higher than non-ICU inpatient costs in these countries.

#### Cost per case in the private settings

Overall, the cost per case in private settings was higher than that in public settings across all treatment settings and countries. The average cost per case in private settings was found to be the highest for direct medical costs for inpatients in Brazil ($1,094.2). For outpatient care, costs in private settings ranged from 1.1 times higher than public settings in India’s direct medical costs to 3.3 times higher in Brazil’s productivity costs (see Appendix 6 in the supplementary file). Similarly, for inpatient care, private hospitals had costs ranging from 1.2 times higher in Indonesia’s direct medical costs to 4.5 times higher in India’s direct medical costs (see Appendix 7 in the supplementary file). India was the only country to report ICU costs in private hospitals, with an average direct medical cost of $901.4.

### Linear regression analysis

The results of the linear regression analysis for outpatients and inpatients treated in public settings are illustrated in Fig. 3. Detailed results, including adjusted R-squared, coefficients, 95% confidence intervals, and p-values, are shown in Appendix 8 and Appendix 9 in the supplementary file for outpatients and inpatients, respectively.

**Fig. 3.**
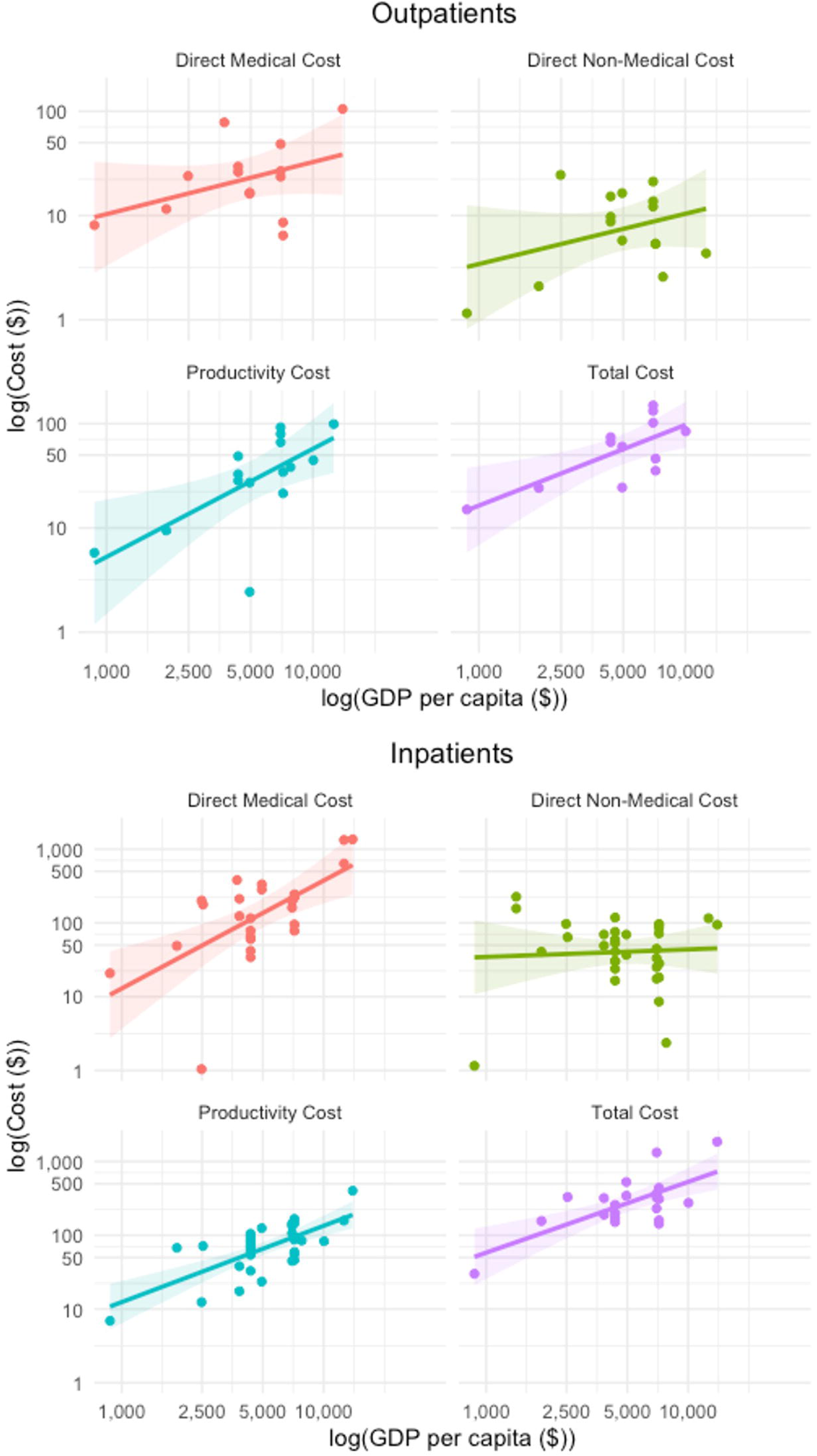
Linear regression analysis between logarithm of cost per case and logarithm of GDP per capita in outpatient and inpatient public settings

#### Shaded area reflects the 95% confidence interval

For outpatients treated in public settings, the logarithm of GDP per capita was positively associated with the logarithm of all cost types. However, this relationship was statistically significant only for productivity costs and total costs (Appendix 8 in the supplementary file). We found that for each unit increase in the GDP per capita on the log scale. The logarithm of total cost increased by 0.7737 GDP (p-value = 0.00762). Similarly, the logarithm of the productivity cost increased by 1.0369 (p = 0.00789). Although the coefficients for other cost types were also positive, these associations were not statistically significant (Appendix 8 in the supplementary file). The adjusted R-squared was highest for the total cost model (0.4784), followed closely by the productivity cost model (0.3868).

For inpatients treated in public settings, the logarithm of GDP per capita was significantly associated not only with the logarithm of productivity costs and of total costs but also with the logarithm of direct medical costs (Appendix 9 in the supplementary file). Each unit increase in logarithm of GDP per capita was associated with a 1.0342 increase in the log of productivity costs (p < 0.001), a 0.9596 increase in the log of total costs (p = 0.0005), and a 1.4632 increase in the log of direct medical costs (p = 0.0005). The association with direct non-medical costs, however, was not statistically significant (Appendix 9 in the supplementary file). Among the significant models, the highest adjusted R-squared was observed for the productivity cost model (0.4796), followed by the total cost model (0.4529) and the direct medical cost model (0.3748).

### Quality of studies

The quality of all the 56 included studies was assessed using the checklist developed by Schnitzler et al. (16). A summary of the results of the quality assessment is shown in Appendix 10 (with more detailed information in Appendix 11 in the supplementary file).

It was found that all included studies performed well in reporting the measurement of resources. While 61% of the studies reported patient characteristics and geographic locations, many lacked clarity regarding disease severity and patients’ comorbidities. Only 39% of the studies clearly described and justified the perspective of the study, whereas 38% of the studies clearly reported the valuation of resources. Furthermore, only some studies (29%) clearly reported the time horizon used. Moreover, only 18% of the studies conducted a sensitivity analysis to estimate uncertainty when projecting the national economic burden of dengue, using the primary cost per case collected.

Apart from the quality of the studies according to the checklist, it was observed that several studies despite referring to productivity costs (or indirect costs) in their methods only reported the number of missed workdays instead (i.e. did not monetise the productivity losses). In addition, several studies had unclear cost categories, resulting in uncertainty around the appropriate category to use to classify them.

## Discussion

This study updates our understanding of the cost of dengue-related illness globally based on a systematic review of the empirical evidence. While the economic burden of dengue was projected for 2013 by Shepard et al. (10), this present study analysed additional primary data and further stratified the sub-types of direct cost analysed. These estimations can serve as inputs for future economic evaluation studies for upcoming dengue interventions (such as vaccine programmes). It should be noted that such economic evaluation studies are an important source of evidence to support decision-makers on the adoption of dengue interventions to alleviate the burden on patients. While the primary data summarised could be directly used for countries where the data are available, the updated regression based projections of costs could be considered for countries that still lack data, but where a timely decision to evaluate programmes is required.

### Cost-of-illness data availability

This review found that dengue cost-of-illness studies are concentrated in the Western Pacific and Southeast Asia. Only one study conducted in Africa, capturing costs in Burkina Faso and Kenya, was identified. It is also evident that real-world cost per case data for dengue remains insufficient in the Eastern Mediterranean and Africa. Additionally, there is a lack of data on the costs for critically ill patients treated in ICU settings in almost every country.

### Factors associated with cost per case variation among regions and countries

The average reported cost per case across countries, stratified by treatment and provider settings, revealed that the cost of dengue illness can be substantial, particularly for patients treated in private hospitals, and even higher for those requiring ICU care. This aligns with the general understanding that treatment and service prices are marked up in private hospitals, while ICU care is more resource-intensive than other treatment settings.

Our results show that, from the included studies, the cost of dengue per case varied across regions, with the highest total costs observed in the Americas, followed by the Western Pacific, Southeast Asia, and the lowest in Africa. Considering the number of high-income (HIC), upper-middle-income (UMIC), lower-middle-income (LMIC), and low-income countries (LIC) included in this study for each region, the Americas region had the highest number of HICs (four countries: French Guiana, Martinique, Guadeloupe, and Panama), whereas the rest were UMICs. On the other hand, the Western Pacific region included one HIC (Japan), two UMICs (China, and Malaysia), and three LMICs (Cambodia, Viet Nam, and the Philippines), while Southeast Asia included two UMICs (Thailand, and Indonesia) and four LMICs (Bangladesh, Cambodia, India, and Sri Lanka). The African region included one LMIC (Kenya) and one LIC (Burkina Faso). Regions with a higher number of HICs and UMICs tend to have higher total costs of dengue, likely due to the higher GDP per capita in those regions. It is crucial to acknowledge that these findings may not be fully representative of the entire region, as the data collection did not encompass all countries within each region.

We also found that GDP per capita was significantly positively associated with the total cost and productivity cost for both outpatients and inpatients treated in public facilities. It was not surprising that indirect costs were associated with GDP per capita, because higher-income countries typically have higher wages. In addition, since some studies use GDP per capita as a proxy for productivity loss, this naturally strengthens the association. The association between GDP per capita and productivity costs also aligns with the studies by Hung et al. (4) and Shepard et al. (10).

There was a positive association between direct non-medical costs and GDP per capita but it was not statistically significant in either group. On the other hand, the direct medical costs were significantly positively associated with GDP per capita for inpatients, but not for outpatients. This might be because outpatient care generally involves fewer medical resources (such as staff time, diagnostic tests, and basic medications) than inpatient care, making the costs less sensitive to a country’s economy. When considering the adjusted R-squared values, the productivity cost provided adjusted R-squared values of 39% and 48% for outpatients and inpatients, respectively. The lower adjusted R-squared found in this study compared to that found in Shepard et al. (10) might be due to the inclusion of more data in this study, resulting in greater variation in the regression model.

### Quality assessment

Overall, the included studies performed well in the results and reporting section, but there is large room for improvement in reporting costs by type, as several studies incorrectly classified costs, and others reported unclear costs, resulting in unusable data in this context. It should be noted that transparently reporting costs by type would benefit future studies, particularly economic evaluations. Nonetheless, the methodology and cost analysis sections of published papers typically require the most improvement, as only a small number of studies reported the epidemiological approach (based on incidence or prevalence data), and the costing and data collection approaches were at times not clearly reported. Although these can be inferred from the study context, it is important to explicitly report them, as inferences can vary depending on readers’ experiences and perspectives. Moreover, the majority of the studies did not clearly report the time horizon used for data collection, whether it included pre-hospitalisation, during hospitalisation, and post-hospitalisation periods. This omission affects the reporting of total costs, as some studies may underestimate total costs if they do not account for the entire illness period, and readers may not be aware of the underestimation if the time horizon is not clearly stated.

### Limitations

This study has several limitations.

First, there was significant variation in methodology among the studies included in the analysis. It is important to note that each study collected data from different patient populations, which had distinct characteristics, such as disease severity and age groups. Although this study reported the cost per case by treatment setting and provider type, we did not analyse the data by finer stratifications of disease severity or age groups due to the limited sample sizes. In addition, while we made our best efforts to compare costs only from studies that used a similar study design —comparing direct medical costs only among studies that reported both government and household direct medical costs or those that reported total costs without stratification, and comparing total costs only among studies that reported all cost sub-types—some variation in the reported costs, due to differences in data collection methods and cost categories (for example, some studies included only treatment costs as direct medical costs, while others also accounted for service fees) remained. Methodological differences in data collection can confound actual differences in costs by country.

Second, we calculated the average cost per case in each country or region by averaging the reported costs, and we did not weight the costs by the number of cases observed, as some studies did in the past, because these did not report the number of study participants.

Furthermore, the time horizon used to capture the costs can affect the total estimated costs but was often not specified. In addition, none of the included studies accounted for the potential burden of dengue-related post-acute consequences, which could also underestimate the cost of illness (71).

Finally, it is important to note that only one researcher performed the screening and data extraction. However, any uncertainties were resolved by discussion with a second reviewer.

### Future research areas

This study provides a valuable deeper understanding of the economic burden of dengue, particularly the variation across settings. Nonetheless, several research gaps still exist and require further investigation in future studies.

To date, insufficient cost-of-illness studies have been conducted in the African and Eastern Mediterranean regions, which limits our understanding of the economic burden on healthcare systems and households in these regions.

Furthermore, a deeper understanding of the drivers of cost variation is crucial for improving cost estimates in future economic evaluations. Notably, only a few countries have reported the cost per case for critically ill patients treated in ICUs. Given that ICU costs were found to be substantially higher than those for other patient groups, more data are needed to assess the economic burden specific to this group. Additionally, further research is required to examine how costs vary by age and the presence of comorbidities.

Some studies also suggest that dengue symptoms could persist for months in some patients, which could significantly affect the economic burden of dengue (71, 72). However, none of the studies included in this systematic review considered these long-term symptoms (71–73). Future studies should explore the potential impact of long-term dengue symptoms to accurately estimate the economic burden of dengue in both child and adult populations.

## Conclusions

The average total cost per case varies significantly across regions and countries. Based on the included studies, the Americas reported the highest average total cost per case, while other regions tended to experience lower costs. Regression analysis revealed that the cost of dengue illness varies significantly across countries and regions, and is positively related to the settings GDP per capita. The results presented in this study can serve as input parameters for future economic evaluations, supporting decision-makers in resource allocation to ensure patient access to dengue interventions.

## Data Availability

All data produced in the present work are contained in the manuscript.

## Declarations

## Ethics approval and consent to participate

Not applicable.

## Consent for publication

Not applicable.

## Availability of data and materials

The authors confirm that the data supporting the findings of this study are available within the article.

## Authors’ contributions

DL performed the systematic review, including study selection, data extraction and adjustment, data analysis, quality assessment, and manuscript writing. ID and HCT designed the study methods, supervised DL, and resolved any uncertainties regarding the inclusion or exclusion of studies. ID and HCT also validated the results, and reviewed and finalised the manuscript. All authors read and approved the final manuscript.

## Acknowledgements

None

