## Supplementary file for "The Economic Burden of Dengue: A Systematic Literature Review of Cost-of-Illness Studies"

**Appendix 1 PRISMA 2020 checklist**

| **Section and Topic** | **Item #** | **Checklist item** | **Location/page where item is reported** |
| --- | --- | --- | --- |
| **TITLE** | | |  |
| Title | 1 | Identify the report as a systematic review. | 1 |
| **ABSTRACT** | | |  |
| Abstract | 2 | See the PRISMA 2020 for Abstracts checklist. | 2 |
| **INTRODUCTION** | | |  |
| Rationale | 3 | Describe the rationale for the review in the context of existing knowledge. | 3-4 |
| Objectives | 4 | Provide an explicit statement of the objective(s) or question(s) the review addresses. | 4 |
| **METHODS** | | |  |
| Eligibility criteria | 5 | Specify the inclusion and exclusion criteria for the review and how studies were grouped for the syntheses. | 4-5 |
| Information sources | 6 | Specify all databases, registers, websites, organisations, reference lists and other sources searched or consulted to identify studies. Specify the date when each source was last searched or consulted. | 4-5 |
| Search strategy | 7 | Present the full search strategies for all databases, registers and websites, including any filters and limits used. | 4-5 |
| Selection process | 8 | Specify the methods used to decide whether a study met the inclusion criteria of the review, including how many reviewers screened each record and each report retrieved, whether they worked independently, and if applicable, details of automation tools used in the process. | 4-5  Appendix 2 |
| Data collection process | 9 | Specify the methods used to collect data from reports, including how many reviewers collected data from each report, whether they worked independently, any processes for obtaining or confirming data from study investigators, and if applicable, details of automation tools used in the process. | 5-7 |
| Data items | 10a | List and define all outcomes for which data were sought. Specify whether all results that were compatible with each outcome domain in each study were sought (e.g. for all measures, time points, analyses), and if not, the methods used to decide which results to collect. | 5-7 |
|  | 10b | List and define all other variables for which data were sought (e.g. participant and intervention characteristics, funding sources). Describe any assumptions made about any missing or unclear information. | 5-7 |
| Study risk of bias assessment | 11 | Specify the methods used to assess risk of bias in the included studies, including details of the tool(s) used, how many reviewers assessed each study and whether they worked independently, and if applicable, details of automation tools used in the process. | 4-5 |
| Effect measures | 12 | Specify for each outcome the effect measure(s) (e.g. risk ratio, mean difference) used in the synthesis or presentation of results. | 5-6 |
| Synthesis methods | 13a | Describe the processes used to decide which studies were eligible for each synthesis (e.g. tabulating the study intervention characteristics and comparing against the planned groups for each synthesis (item #5)). | 6-7 |
|  | 13b | Describe any methods required to prepare the data for presentation or synthesis, such as handling of missing summary statistics, or data conversions. | 6-7 |
|  | 13c | Describe any methods used to tabulate or visually display results of individual studies and syntheses. | 6-7 |
|  | 13d | Describe any methods used to synthesize results and provide a rationale for the choice(s). If meta-analysis was performed, describe the model(s), method(s) to identify the presence and extent of statistical heterogeneity, and software package(s) used. | 6-7 |
|  | 13e | Describe any methods used to explore possible causes of heterogeneity among study results (e.g. subgroup analysis, meta-regression). | 6-7 |
|  | 13f | Describe any sensitivity analyses conducted to assess robustness of the synthesized results. | No |
| Reporting bias assessment | 14 | Describe any methods used to assess risk of bias due to missing results in a synthesis (arising from reporting biases). | No |
| Certainty assessment | 15 | Describe any methods used to assess certainty (or confidence) in the body of evidence for an outcome. | No |
| **RESULTS** | | |  |
| Study selection | 16a | Describe the results of the search and selection process, from the number of records identified in the search to the number of studies included in the review, ideally using a flow diagram. | 8-9 |
|  | 16b | Cite studies that might appear to meet the inclusion criteria, but which were excluded, and explain why they were excluded. | No |
| Study characteristics | 17 | Cite each included study and present its characteristics. | 8-10 |
| Risk of bias in studies | 18 | Present assessments of risk of bias for each included study. | No |
| Results of individual studies | 19 | For all outcomes, present, for each study: (a) summary statistics for each group (where appropriate) and (b) an effect estimate and its precision (e.g. confidence/credible interval), ideally using structured tables or plots. | 8-10,  Appendix 4 |
| Results of syntheses | 20a | For each synthesis, briefly summarise the characteristics and risk of bias among contributing studies. | 10-15 |
|  | 20b | Present results of all statistical syntheses conducted. If meta-analysis was done, present for each the summary estimate and its precision (e.g. confidence/credible interval) and measures of statistical heterogeneity. If comparing groups, describe the direction of the effect. | 10-15 |
|  | 20c | Present results of all investigations of possible causes of heterogeneity among study results. | 8-10, 14-15 |
|  | 20d | Present results of all sensitivity analyses conducted to assess the robustness of the synthesized results. | No |
| Reporting biases | 21 | Present assessments of risk of bias due to missing results (arising from reporting biases) for each synthesis assessed. | No |
| Certainty of evidence | 22 | Present assessments of certainty (or confidence) in the body of evidence for each outcome assessed. | No |
| **DISCUSSION** | | |  |
| Discussion | 23a | Provide a general interpretation of the results in the context of other evidence. | 15-18 |
|  | 23b | Discuss any limitations of the evidence included in the review. | 18-19 |
|  | 23c | Discuss any limitations of the review processes used. | 18-19 |
|  | 23d | Discuss implications of the results for practice, policy, and future research. | 19-20 |
| **OTHER INFORMATION** | | |  |
| Registration and protocol | 24a | Provide registration information for the review, including register name and registration number, or state that the review was not registered. | No |
|  | 24b | Indicate where the review protocol can be accessed, or state that a protocol was not prepared. | No |
|  | 24c | Describe and explain any amendments to information provided at registration or in the protocol. | No |
| Support | 25 | Describe sources of financial or non-financial support for the review, and the role of the funders or sponsors in the review. | No |
| Competing interests | 26 | Declare any competing interests of review authors. | No |
| Availability of data, code and other materials | 27 | Report which of the following are publicly available and where they can be found: template data collection forms; data extracted from included studies; data used for all analyses; analytic code; any other materials used in the review. | 9-10, Fig 1, Appendix 2, Appendix 4 |

**Reference:** (1)

**Appendix 2 Details of keyword used in each database.**

| **Database** | **Full keywords** |
| --- | --- |
| MEDLINE via Ovid &  EMBASE via Ovid | 1 exp Dengue/ or exp Severe Dengue/ or exp Dengue Virus/  2 dengue.mp. [mp=title, book title, abstract, original title, name of substance word, subject heading word, floating sub-heading word, keyword heading word, organism supplementary concept word, protocol supplementary concept word, rare disease supplementary concept word, unique identifier, synonyms, population supplementary concept word, anatomy supplementary concept word]  3 DENV.mp. [mp=title, book title, abstract, original title, name of substance word, subject heading word, floating sub-heading word, keyword heading word, organism supplementary concept word, protocol supplementary concept word, rare disease supplementary concept word, unique identifier, synonyms, population supplementary concept word, anatomy supplementary concept word]  4 DENV-1.mp. [mp=title, book title, abstract, original title, name of substance word, subject heading word, floating sub-heading word, keyword heading word, organism supplementary concept word, protocol supplementary concept word, rare disease supplementary concept word, unique identifier, synonyms, population supplementary concept word, anatomy supplementary concept word]  5 DENV-2.mp. [mp=title, book title, abstract, original title, name of substance word, subject heading word, floating sub-heading word, keyword heading word, organism supplementary concept word, protocol supplementary concept word, rare disease supplementary concept word, unique identifier, synonyms, population supplementary concept word, anatomy supplementary concept word]  6 DENV-3.mp. [mp=title, book title, abstract, original title, name of substance word, subject heading word, floating sub-heading word, keyword heading word, organism supplementary concept word, protocol supplementary concept word, rare disease supplementary concept word, unique identifier, synonyms, population supplementary concept word, anatomy supplementary concept word]  7 DENV-4.mp. [mp=title, book title, abstract, original title, name of substance word, subject heading word, floating sub-heading word, keyword heading word, organism supplementary concept word, protocol supplementary concept word, rare disease supplementary concept word, unique identifier, synonyms, population supplementary concept word, anatomy supplementary concept word] 581  8 1 or 2 or 3 or 4 or 5 or 6 or 7  9 exp "Cost of Illness"/ or exp "Costs and Cost Analysis"/  10 exp Financial Stress/  11 exp Health Care Costs/  12 cost of illness.mp. [mp=title, book title, abstract, original title, name of substance word, subject heading word, floating sub-heading word, keyword heading word, organism supplementary concept word, protocol supplementary concept word, rare disease supplementary concept word, unique identifier, synonyms, population supplementary concept word, anatomy supplementary concept word] 34138  13 economic burden.mp. [mp=title, book title, abstract, original title, name of substance word, subject heading word, floating sub-heading word, keyword heading word, organism supplementary concept word, protocol supplementary concept word, rare disease supplementary concept word, unique identifier, synonyms, population supplementary concept word, anatomy supplementary concept word]  14 9 or 10 or 11 or 12 or 13  15 8 and 14 |
| Web of Science | (ALL=(dengue)) AND ALL=(cost of illness OR economic burden) |
| PubMed | (dengue) AND (cost of illness OR economic burden) |

**Appendix 3 Assumptions for data analysis.**

Since the World Bank’s exchange rate from Sri Lankan Rupees and Venezuela Bolívar to USD for 2023 were unavailable, the exchange rate from the latest available year was used. Furthermore, for French Guiana, Martinique, Guadeloupe, and Venezuela, where the GDP deflator from the World Bank was not available, the inflation to 2023 values was adjusted using US inflation rates, as this was recommended for countries where GDP deflator data was not available (2). Although WHO classified French Guiana, Martinique, and Guadeloupe as an ‘unknown’ region, we grouped them into the Americas region based on geographic location to facilitate analysis. Finally, in the linear regression analysis, since GDP per capita data for these French territories and Venezuela were not available from the World Bank, we sourced the latest figures from alternative sources. The latest GDP per capita for these French territories were extracted from the Pan America Health Organisation report, while for Venezuela, the GDP per capita in 2023 from the International Monetary Fund was used (3).

**Appendix 4 Details of included studies**

| WHO region | Country | Publication year | Perspective | Provider setting | Treatment setting | Data collection approach | Costing approach | Dengue severity | Age group | Study |
| --- | --- | --- | --- | --- | --- | --- | --- | --- | --- | --- |
| African & Western Pacific | Burkina Faso, Kenya, Cambodia | 2019 | Societal | Public | Outpatient, Inpatient | Prospective | Bottom-up | Unclear | Children and adult* | Lee et al. (4) |
| Americas & Soth-East Asia & Western Pacific | Viet Nam, Thailand, Colombia | 2017 | Societal | Public | Outpatient,  Inpatient | Prospective | Bottom-up | DF and DHF/DSS* | Children, adult | Lee et al. (5) |
| Americas & Soth-East Asia & Western Pacific | Brazil, El Salvador, Guatemala, Panama, Venezuela, Cambodia, Malaysia, Thailand | 2009 | Societal | Public and private* | Outpatient, Inpatient | Prospective | Macro-costing | DF and DHF/DSS* | Children and adult* | Suaya et al. (6) |
| Americas | Brazil | 2014 | Government & private | Public,  Private | Inpatient and ICU* | Retrospective | Bottom-up | DF and DHF/DSS* | Children and adult* | Vieira et al. (7) |
| Americas | Brazil | 2015 | Societal | Public, Private | Outpatient, inpatient | Prospective | Micro-costing | DF and DHF/DSS* | Children and adult* | Martelli et al. (8) |
| Americas | Brazil | 2018 | Government | Public | Inpatient | Retrospective | Unclear | DF, DHF/DSS | Children and adult* | Godói et al. (9) |
| Americas | Brazil | 2022 | Private health insurance | Private | Inpatient | Retrospective | Bottom-up | DF and DHF/DSS* | Children and adult* | Abud et al. (10) |
| Americas | Colombia | 2015 | Societal | Public | Outpatient, Inpatient | Retrospective | Bottom-up | DF, DHF/DSS | Children and adult* | Castro Rodriguez et al. (11) |
| Americas | French Guiana, Martinique, Guadeloupe | 2016 | Government | Public | Inpatient and ICU* | Retrospective | Bottom-up | Unclear | Children and adult* | Uhart et al. (12) |
| Americas | Mexico | 2016 | Societal** | Public | Outpatient,  Inpatient,  Intensive care unit | Retrospective | Micro-costing | Unclear | Children and adult* | Zubieta-Zavala et al. (13) |
| Americas | Mexico | 2016 | Government | Public | Inpatient,  Intensive care unit | Retrospective | Gross-costing + activity-based | DF, DHF/DSS | Children, adult | Thalagala et al. (14) |
| Americas | Mexico | 2017 | Household | Public, Private | Outpatient, Inpatient, Informal care | Retrospective | Bottom-up | DF and DHF/DSS* | Children and adult* | Legorreta-Soberanis et al. (15) |
| Americas | Panama | 2008 | Societal | Public and private* | Outpatient and inpatient* | Prospective | Bottom-up + Macro-costing | DF and DHF/DSS* | Children, adult | Armien et al. (16) |
| Americas | Peru | 2015 | Household | Public | Outpatient, Inpatient | Retrospective | Bottom-up | DF and DHF/DSS* | Children and adult* | Salmon-Mulanovich et al. (17) |
| Eastern mediterranean | Pakistan | 2009 | Unclear | Private | Inpatient | Prospective | Bottom-up | DF | Adult | Riaz et al. (18) |
| Eastern mediterranean | Pakistan | 2011 | Household | Public | Inpatient | Retrospective | Bottom-up | DF and DHF/DSS* | Children and adult* | Rafique et al. (19) |
| Eastern Mediterranean | Pakistan | 2019 | Household | Public | Inpatient | Prospective | Bottom-up | DF and DHF/DSS* | Children and adult* | Jamil et al. (20) |
| Eastern Mediterranean | Saudi Arabia | 2020 | Societal | Public and private* | Outpatient and inpatient* | Retrospective | Bottom-up | DF and DHF/DSS* | Children and adult* | Akbar et al. (21) |
| South-east Asia | Bangladesh | 2023 | Societal | Public, Private | Inpatient | Retrospective & Prospective | Micro-costing | Unclear | Children and adult* | Sarker et al. (22) |
| South-east Asia | India | 2008 | Unclear | Private | Inpatient, ICU | Unclear | Bottom-up | DF and DHF/DSS* | Children and adult* | Garg et al. (23) |
| South-east Asia | India | 2014 | Societal** | Public,  Private | Outpatient, Inpatient | Retrospective&  Prospective | Macro-costing | Unclear | Children and adult* | Shepard et al. (24) |
| South-east Asia | India | 2015 | Societal | Private | Inpatient | Prospective | Bottom-up | DF and DHF/DSS* | Children | Manjunath et al. (25) |
| South-east Asia | India | 2019 | Household | Private | Inpatient | Retrospective | Bottom-up | DF, DHF/DSS | Children, adult | Panmei et al. (26) |
| South-east Asia | India | 2019 | Societal** | Public, Private | Inpatient | Retrospective | Bottom-up | DF and DHF/DSS* | Children and adult* | Bajwala et al. (27) |
| South-east Asia | India | 2020 | Household | Public | Outpatient, inpatient | Retrospective | Bottom-up | Unclear | Children and adult* | Nujum et al. (28) |
| South-east Asia | India | 2021 | Unclear | Public and private* | Inpatient | Prospective | Unclear | Unclear | Children | Srinivasan et al. (29) |
| South-east Asia | India | 2021 | Household | Public | Inpatient | Retrospective | Bottom-up | Unclear | Adult | Rafikahmed et al. (30) |
| South-east Asia | India | 2022 | Household | Public, Private | Outpatient and inpatient* | Prospective | Bottom-up | Unclear | Children and adult* | Kaur et al. (31) |
| South-east Asia | Indonesia | 2019 | Household | Public | Inpatient | Retrospective | Bottom-up | DHF/DSS | Children and adult* | Supadmi et al. (32) |
| South-east Asia | Indonesia | 2019 | Societal | Private, Public | Outpatient, Inpatient | Retrospective & Prospective | Bottom-up | DF and DHF/DSS* | Children and adult* | Nadjib et al. (33) |
| South-east Asia | Indonesia | 2020 | Unclear | Public | Outpatient, Inpatient,  Informal care | Prospective | Macro-costing | Unclear | Children and adult* | Wilastonegoro et al. (34) |
| South-East Asia | Sri Lanka | 2021 | Societal | Public | Inpatient | Prospective | Bottom-up | DF, DHF/DSS | Children | Sonali Fernando et al. (35) |
| South-East Asia | Sri Lanka | 2021 | Govern-ment | Public | Inpatient | Prospective | Bottom-up | DF, DHF/DSS | Children and adult* | Sigera et al. (36) |
| South-east Asia | Sri Lanka | 2022 | Societal | Public | Inpatient and ICU* | Prospective | Gross-costing &  Activity-based | DF and DHF/DSS* | Children and adult* | Weerasinghe et al. (37) |
| South-east Asia | Thailand | 1997 | Societal | Public | Inpatient | Prospective | Bottom-up | DHF/DSS | Children, adult | Okanurak et al. (38) |
| South-east Asia | Thailand | 2005 | Household | Public | Inpatient | Retrospective | Bottom-up | DF and DHF/DSS* | Children and adult* | Clark et al. (39) |
| South-east Asia | Thailand | 2017 | Household | Public | Inpatient | Prospective | Bottom-up | DF, DHF/DSS | Children, adult | Tozan et al. (40) |
| Western pacific | Cambodia | 2004 | Household | Public and private | Inpatient and ICU* | Retrospective | Bottom-up | Unclear | Unclear | Van Damme et al. (41) |
| Western pacific | Cambodia | 2008 | Household | Public, Private | Inpatient | Prospective | Bottom-up | Unclear | Children | Khun et al. (42) |
| Western pacific | Cambodia | 2009 | Household | Public and private* | Outpatient and inpatient* | Prospective | Bottom-up | Unclear | Children | Huy et al. (43) |
| Western pacific | China | 2017 | Government | Public | Inpatient | Retrospective | Gross-costing | DF and DHF/DSS* | Children, adult | Zhang et al. (44) |
| Western pacific | China | 2022 | Societal | Public | Outpatient and inpatient* | Retrospective | Bottom-up | DF and DHF/DSS* | Children and adult* | Xu et al. (45) |
| Western pacific | China | 2022 | Unclear | Public | Inpatient | Retrospective | Bottom-up | DF | Children | Wang et al. (46) |
| Western pacific | China | 2023 | Household | Public | Outpatient, Inpatient | Retrospective | Bottom-up | DF and DHF/DSS* | Children and adult* | Yu et al. (47) |
| Western pacific | Japan | 2020 | Societal** | Public | Outpatient, inpatient | Retrospective | Bottom-up | DF, DHF/DSS | Children and adult* | Kajimoto et al. (48) |
| Western pacific | Malaysia | 2016 | Household | Public | Outpatient and inpatient* | Retrospective | Bottom-up | Unclear | Children and adult* | Mia et al. (49) |
| Western pacific | Philippines | 2015 | Societal** | Public, Private | Outpatient, Inpatient | Retrospective | Macro-costing | DF and DHF/DSS* | Unclear | Edillo et al. (50) |
| Western pacific | Philippines | 2016 | Societal** | Public and private* | Inpatient | Retrospective | Bottom-up | Unclear | Children, adult | Onuh et al. (51) |
| Western pacific | Viet Nam | 2007 | Household | Public | Inpatient | Unclear | Bottom-up | DHF/DSS | Children | Harving et al. (52) |
| Western pacific | Viet Nam | 2012 | Household | Public | Inpatient | Retrospective | Bottom-up | Unclear | Children and adult* | Tam et al. (53) |
| Western pacific | Viet Nam | 2016 | Household | Public | Inpatient | Prospective | Bottom-up | DHF/DSS | Children, adult | Nhi et al. (54) |
| Western pacific | Viet Nam | 2017 | Unclear | Public | Inpatient | Retrospective | Bottom-up | DF and DHF/DSS* | Children, adult | Vo et al. (55) |
| Western pacific | Viet Nam | 2017 | Societal | Public | Inpatient | Retrospective | Bottom-up | Unclear | Children, adult | Pham et al. (56) |
| Western pacific | Viet Nam | 2018 | Household | Public | Outpatient | Retrospective | Bottom-up | DF and DHF/DSS* | Unclear | Tran et al. (57) |
| Western pacific | Viet Nam | 2019 | Societal | Public | Inpatient and ICU* | Prospective | Bottom-up | DHF/DSS | Adult | McBride et al. (58) |
| Western pacific | Viet Nam | 2022 | Societal | Public | ICU patient | Retrospective | Bottom-up | Unclear | Adult | Hung et al. (59) |

Note: * indicates not stratify each sub-group, ** using societal perspective but did not report all cost types.

Abbreviation: WHO, World Health Organization; DF, dengue fever; DHF, dengue haemorrhagic fever; DSS, dengue shock syndrome; ICU, intensive care unit.

**Appendix … Comparison of cost per case between private and public hospitals across countries (2023 USD).**

**Appendix 5 Average reported cost per case in outpatient private setting across countries (2023 USD).**

|  | Average total cost | Average direct medical cost | Average direct non-medical cost | Average productivity cost |
| --- | --- | --- | --- | --- |
| Americas |  |  |  |  |
| Brazil | 252.64 |  |  | 147.64 |
| South-east Asia | |  |  |  |
| India |  | 26.58 |  |  |
| Indonesia | 86.52 | 33.96 | 18.98 | 33.59 |
| Western pacific | |  |  |  |
| Philippines |  | 165.68 |  |  |

**Appendix 6 Average reported cost per case in inpatient private setting across countries (2023 USD)**

|  | Average total cost | Average direct medical cost | Average direct non-medical cost | Average productivity cost |
| --- | --- | --- | --- | --- |
| Americas |  |  |  |  |
| Brazil |  | 1094.23 |  | 219.54 |
| Eastern mediterranean | |  |  |  |
| Pakistan |  | 518.60 |  |  |
| South-east Asia | |  |  |  |
| Bangladesh | 546.88 | 341.59 | 91.24 | 103.63 |
| India |  | 449.59 |  | 35.94 |
| Indonesia | 537.94 | 359.47 | 77.17 | 101.30 |

**Appendix 7 Result of the linear regression in outpatient public hospitals.**

| Cost type | Number of studies  included | Adjusted R-square | Coefficient | 95% CI Lower  (2.5%) | 95% CI Upper  (97.5%) | Intercept | P-value |
| --- | --- | --- | --- | --- | --- | --- | --- |
| Direct medical cost | 14 | 0.10 | 0.50 | -0.19 | 1.19 | -1.12 | 0.14 |
| Direct non-medical cost | 15 | 0.06 | 0.48 | -0.27 | 1.24 | -2.10 | 0.19 |
| Indirect cost | 15 | 0.39 | 1.04 | 0.32 | 1.75 | -5.50 | 0.008 ** |
| Total cost | 12 | 0.48 | 0.77 | 0.26 | 1.30 | -2.55 | 0.008 ** |

Note: * indicates statistically significant; ** indicates a strong statistically significant.

**Appendix 8 Result of the linear regression in inpatient public hospitals.**

| Cost type | Number of studies included | Adjusted R-square | Coefficient | 95% CI Lower  (2.5%) | 95% CI Upper  (97.5%) | Intercept | P-value |
| --- | --- | --- | --- | --- | --- | --- | --- |
| Direct medical cost | 26 | 0.37 | 1.46 | 0.71 | 2.22 | -7.55 | 0.0005 ** |
| Direct non-medical cost | 34 | -0.03 | 0.10 | -0.54 | 0.75 | 2.84 | 0.75 |
| Indirect cost | 33 | 0.48 | 1.03 | 0.65 | 1.42 | -4.62 | 4.81e-06*** |
| Total cost | 21 | 0.45 | 0.96 | 0.48 | 1.44 | -2.57 | 0.0005 ** |

Note: * indicates statistically significant; ** indicates a strong statistically significant; *** indicates a very strong statistically significant.

**Appendix 9 Full result of the quality assessment of the included studies.**

| **Study** | **Question/ objective** | **Population** | **Perspective** | **Epidemiology approach** | **Costing approach** | **Data collection** | **Identification of resource** | **Measurement of resource** | **Valuation of resource** | **Time horizon** | **Discounting** | **Sensitivity** | **Cost sectors** | **Generalizability** | **Limitations** | **Ethical and distributional issues** | **Conflict of interest** |
| --- | --- | --- | --- | --- | --- | --- | --- | --- | --- | --- | --- | --- | --- | --- | --- | --- | --- |
| **Yu et al. (47)** | Yes | Yes | Partial | **No** | Unclear | Yes | Yes | Yes | Partial | **No** | NA | No | Yes | Yes | Yes | No | Yes |
| **Sarker et al. (22)** | Yes | Yes | Yes | Yes | Yes | Unclear | Yes | Yes | Partial | Partial | NA | Partial | Yes | Yes | Yes | Yes | Yes |
| **Xu et al. (45)** | Yes | Partial | Partial | No | Unclear | Yes | Yes | Yes | Partial | No | NA | No | Yes | Yes | Yes | No | Yes |
| **Weerasinghe et al. (37)** | Yes | Yes | Yes | No | Yes | Yes | Yes | Yes | Partial | Partial | No | No | Yes | Yes | Yes | No | Yes |
| **Wang et al. (46)** | Partial | Yes | No | Yes | Unclear | Yes | No | Yes | No | Yes | No | No | No | No | Yes | No | Yes |
| **Kaur et al. (31)** | Partial | Partial | Unclear | No | Unclear | Yes | Yes | Yes | Unclear | Unclear | No | No | Yes | Partial | No | No | Yes |
| **Hung et al. (59)** | Yes | Yes | Yes | No | Yes | Unclear | Yes | Yes | Yes | Yes | No | No | Yes | Yes | Yes | Yes | Yes |
| **Abud et al. (10)** | Partial | Partial | Yes | No | Unclear | Yes | Yes | Yes | Yes | Partial | No | No | No | Yes | Yes | No | Yes |
| **Srinivasan et al. (29)** | Partial | Partial | No | No | No | Yes | Yes | Yes | No | Yes | No | No | No | Yes | Unclear | No | Yes |
| **Sonali Fernando et al. (35)** | Partial | Yes | Partial | No | Unclear | Yes | Yes | Yes | Yes | Yes | NA | No | Yes | Yes | Yes | Unclear | No |
| **Sigera et al. (36)** | Partial | Yes | Yes | No | Unclear | Yes | Yes | Yes | Partial | Partial | No | No | Partial | Yes | Yes | No | Yes |
| **Rafikahmed et al. (30)** | Partial | Partial | Unclear | No | Unclear | Yes | Yes | Yes | Partial | Partial | No | No | Yes | Yes | No | No | Yes |
| **Wilastonegoro, et al (34)** | Partial | Partial | Unclear | No | Yes | Yes | Yes | Yes | Yes | Unclear | NA | No | No | Yes | Yes | No | Yes |
| **Nujum et al. (28)** | Yes | Partial | Yes | No | Unclear | Unclear | Yes | Yes | Yes | Partial | NA | No | Yes | Yes | Unclear | No | Yes |
| **Kajimoto et al. (48)** | Partial | Partial | Yes | No | Unclear | Unclear | No | Yes | Partial | Partial | No | Yes | No | Yes | Yes | No | Yes |
| **Akbar et al. (21)** | Partial | Yes | Unclear | No | Yes | Yes | Yes | Yes | Partial | Unclear | No | No | Yes | Yes | Yes | No | Yes |
| **Supadmi et al. (32)** | Partial | Yes | Unclear | No | No | Yes | Yes | Yes | Partial | Unclear | No | No | No | Partial | No | Partial | Yes |
| **Panmei et al. (26)** | Partial | Yes | Unclear | No | Unclear | Unclear | Yes | Yes | Partial | No | No | No | Yes | Yes | Yes | No | Yes |
| **Nadjib et al. (33)** | Partial | Yes | Yes | No | Unclear | Yes | Partial | Yes | Partial | No | No | Yes | Yes | Yes | Yes | No | Yes |
| **McBride et al. (58)** | Partial | Yes | Unclear | No | Unclear | Yes | No | Yes | Partial | No | No | No | No | Yes | Yes | Yes | Yes |
| **Lee et al. (60)** | Yes | Yes | Yes | No | Unclear | Unclear | Yes | Yes | Partial | No | No | No | Partial | Yes | Yes | Yes | Yes |
| **Jamil et al. (20)** | Yes | Partial | Yes | No | Unclear | Unclear | Yes | Yes | Partial | Yes | No | No | Yes | Partial | No | No | Yes |
| **Bajwala et al. (27)** | Partial | Yes | Partial | Yes | Unclear | Yes | Yes | Yes | Yes | Yes | No | No | Yes | Yes | Yes | Yes | Yes |
| **Tran et al. (57)** | Partial | Yes | Unclear | No | Unclear | Yes | Yes | Yes | No | No | NA | No | Yes | Yes | Yes | No | Yes |
| **Godói et al. (61)** | Yes | Yes | Yes | No | No | Unclear | Partial | Yes | Partial | Partial | No | No | No | Yes | Yes | No | Yes |
| **Zhang et al. (44)** | Partial | Yes | Yes | No | Unclear | Unclear | Yes | Yes | Yes | Yes | No | No | Yes | Yes | Yes | No | Yes |
| **Vo et al. (55)** | Partial | Yes | No | No | Unclear | Yes | Yes | Yes | Partial | Partial | No | No | Yes | Yes | Unclear | No | No |
| **Tozan et al. (40)** | Yes | Yes | Yes | No | Unclear | Yes | Yes | Yes | Partial | Unclear | No | No | Partial | Yes | Yes | No | Yes |
| **Pham et al. (56)** | Yes | Partial | Partial | Yes | Unclear | Yes | Yes | Yes | Partial | Partial | No | Yes | Yes | Yes | Yes | Yes | Yes |
| **Legorreta-Soberanis et al. (15)** | Yes | Yes | Partial | No | Unclear | Unclear | Yes | Yes | Yes | Partial | No | No | No | No | Yes | Yes | Yes |
| **Lee et al. (60)** | Partial | Partial | Unclear | No | Unclear | Unclear | Yes | Yes | Yes | Yes | No | No | Partial | Yes | Yes | No | Yes |
| **Zubieta-Zavala et al. (13)** | Partial | Partial | Yes | No | Yes | Unclear | Yes | Yes | Yes | No | NA | No | Yes | Yes | Yes | No | Yes |
| **Uhart et al. (12)** | Partial | No | Partial | No | Unclear | Unclear | Yes | Yes | Partial | Yes | No | No | No | Yes | Yes | No | Yes |
| **Thalagala et al. (14)** | Yes | Partial | Yes | No | Yes | Yes | Yes | Yes | Yes | Yes | NA | No | Yes | Yes | Yes | No | Yes |
| **Onuh et al. (51)** | Partial | Partial | Unclear | No | Unclear | Unclear | Partial | Yes | Yes | No | No | No | Partial | Yes | Yes | No | Yes |
| **Nhi et al. (54)** | Partial | Yes | Unclear | No | Unclear | Unclear | Yes | Yes | Yes | Yes | No | No | Yes | Yes | Yes | No | Yes |
| **Mia et al. (49)** | Partial | Partial | Yes | No | Unclear | Yes | Yes | Yes | Yes | Yes | No | No | Yes | Yes | Yes | No | No |
| **Salmon-Mulanovich et al. (17)** | Yes | Yes | Yes | No | Unclear | Unclear | Yes | Yes | Yes | No | No | No | Partial | Yes | Yes | Yes | No |
| **Rafique et al. (19)** | Partial | Yes | Yes | No | Unclear | Unclear | Yes | Yes | No | Yes | No | No | Partial | Yes | Yes | No | No |
| **Martelli et al. (8)** | Yes | Yes | Yes | No | Yes | Yes | Yes | Yes | Yes | Partial | No | Yes | Partial | Yes | Yes | No | Yes |
| **Manjunath et al. (25)** | Partial | Yes | Unclear | No | Unclear | Unclear | No | Yes | No | No | NA | No | Partial | Partial | No | No | Yes |
| **Castro Rodriguez et al. (11)** | Partial | Yes | Yes | No | Unclear | Unclear | Yes | Yes | Yes | No | Partial | Yes | Yes | Yes | Yes | No | No |
| **Edillo et al. (50)** | Partial | Partial | No | No | Unclear | Unclear | Yes | Yes | Partial | No | No | Yes | Yes | Yes | Yes | No | No |
| **Vieira et al. (7)** | Partial | Yes | Partial | No | Yes | Yes | Yes | Yes | Yes | No | NA | No | Yes | Yes | Yes | Yes | Yes |
| **Shepard et al. (24)** | Partial | Partial | No | No | Yes | Yes | Yes | Yes | Yes | Yes | No | Yes | Partial | Yes | Yes | No | No |
| **Tam et al. (53)** | Partial | Partial | Unclear | No | Unclear | Unclear | No | Yes | No | Partial | No | No | Yes | Yes | No | Yes | Yes |
| **Suaya et al. (6)** | Partial | Yes | Yes | No | Yes | Yes | Yes | Yes | Yes | Yes | Partial | No | Partial | Yes | Yes | No | Yes |
| **Riaz et al. (18)** | Partial | Yes | No | No | Unclear | Unclear | Partial | Yes | No | No | NA | No | No | No | No | No | No |
| **Huy et al. (43)** | Yes | Partial | Unclear | No | Unclear | Yes | Yes | Yes | Yes | No | NA | No | Partial | Yes | Yes | Yes | Yes |
| **Khun et al. (42)** | Unclear | Yes | Unclear | No | Unclear | Unclear | Yes | Yes | No | No | No | No | No | Yes | No | Yes | Yes |
| **Garga et al. (23)** | Partial | Yes | No | No | Unclear | No | Yes | Yes | No | No | NA | Yes | No | Yes | Yes | No | Yes |
| **Armien et al. (16)** | Partial | Yes | Yes | No | Yes | Yes | Partial | Yes | No | No | Partial | Yes | Partial | No | Yes | No | No |
| **Harving et al. (52)** | Partial | Yes | No | No | Unclear | No | Yes | Yes | No | No | NA | No | Yes | Yes | Yes | No | No |
| **Clark et al. (39)** | Partial | Partial | Unclear | No | Unclear | Unclear | Partial | Yes | No | Yes | NA | Yes | No | Yes | No | No | No |
| **Van Damme et al. (41)** | Unclear | No | Unclear | No | Unclear | Unclear | No | Yes | No | No | NA | No | No | Yes | Yes | Yes | No |
| **Okanurak et al. (38)** | Partial | Yes | Unclear | No | Unclear | Unclear | Partial | Yes | No | Partial | NA | No | Yes | No | Unclear | Yes | No |
